# Mortality in people with attention-deficit/hyperactivity disorder (ADHD): Examining how risk is embodied in a pooling of two prospective cohort studies

**DOI:** 10.64898/2026.06.08.26355148

**Authors:** Haibin Li, Tamsin Ford, Varun Warrier, Steven Bell, G. David Batty

## Abstract

**Background:** Nascent findings suggest that people with attention-deficit/hyperactivity disorder (ADHD) experience higher rates of mortality. To date, study samples have been insufficiently well-characterized to examine the mechanisms via which this neurodevelopmental condition elevates mortality risk.

**Methods:** We used data from the 2007 and 2011 waves of the US National Health Interview Survey, a general population-based cohort study comprising 52097 adults (28675 women) aged 18 years or older at baseline. ADHD diagnosis and an array of demographic, socioeconomic, lifestyle, and co-morbidity (somatic and psychiatric) covariates were self-reported.

**Findings:** At baseline, compared with unaffected individuals, participants with ADHD were more likely to be socioeconomically disadvantaged, smoke cigarettes, consume alcohol, and report symptoms of psychological distress. A median 7.75 years of mortality surveillance (range: 7.25-12.25) gave rise to 6597 deaths from all-causes. After adjustment for age, sex, ethnicity, and survey year, ADHD was associated with a markedly elevated risk of death (hazard ratio [95% confidence interval]: 1.58 [1.20-2.09]). Statistical adjustment for socioeconomic circumstances (11% attenuation), physical co-morbidities (15%), and lifestyle factors (17%) had only a modest impact on the ADHD–death gradient, with the greatest explanatory power apparent for symptoms of depression and anxiety (58%). The magnitude of the association of ADHD with mortality was commensurate to that for several well-established risk factors such as poverty (1.66 [1.55-1.78]), hypertension (1.41 [1.32-1.51]), and diabetes (1.71 [1.59-1.85]) but somewhat lower than cigarette smoking (2.51 [2.29-2.76]) after controlling for age, sex, ethnicity, and survey year. Associations between ADHD and cause-specific mortality from cardiovascular disease, cancer, and chronic respiratory disease were inconclusive.

**Interpretation:** In the present study, the influence of ADHD on total mortality appears to be largely embodied via a series of malleable characteristics, particularly mental illness. If confirmed elsewhere, these results raise the possibility that risk factor modification via standard pharmacological and behavioral interventions could help reduce rates of premature mortality in this patient group.

**Funding:** This paper received no direct funding. GDB is supported by the UK Medical Research Council (MR/P023444/1) and the US National Institute on Aging (1R56AG052519-01, 1R01AG052519-01A1).

**Research in context:** *Evidence before this study:* We searched the PubMed database (January 1947 to June 2026) using the terms ‘attention-deficit/hyperactivity disorder ‘, ‘ADHD’, ‘mortality’, and ‘death’ with no language or date restrictions. We considered individual studies, systematic reviews, and meta-analyses. The most recent systematic review was published in 2022 and the majority of the eight non-overlapping studies found that ADHD was associated with a raised risk of mortality. Included cohort studies were typically generated from linked administrative health records and therefore insufficiently well characterized to comprehensively test how ADHD was embodied in order to elevate death rates. There was some suggestion of a role for mediation via socioeconomic position and mental illness but lifestyle factors such as obesity, smoking, physical inactivity, and alcohol intake had not been considered. A side-by-side comparison of the magnitude of the relationship of ADHD with conventional risk factors for mortality was also lacking.

*Added value of this study:* This is the first prospective cohort study to systematically and comprehensively test the mechanisms via which ADHD elevates mortality and to contextualize the strength of this relationship against well-established risk factors. Socioeconomic circumstances, physical co-morbidities, and lifestyle factors mediated less than one fifth of the ADHD–death gradient, with the greatest explanatory power apparent for depression and anxiety. The impact of ADHD on mortality risk was commensurate to that apparent for several well-established risk factors for mortality such as poverty, hypertension, and diabetes but somewhat lower than estimates for cigarette smoking and physical inactivity.

*Implications of all the available evidence:* While supporting evidence is required, our findings point to the possibility that modification of physical co-morbidities, lifestyle factors, and mental illness via pharmacological treatment and behavioral intervention may have utility in the avoidance of premature mortality in people with ADHD.

## Introduction

A series of studies conducted over several decades has found that, relative to unaffected individuals, there is a higher burden of mortality in people diagnosed with selected psychiatric conditions, including depression, schizophrenia, and personality disorder.^1-4^ Most recently, neurodevelopmental disorders have been examined in this context, with nascent evidence suggesting similarly elevated rates of premature mortality in adults with an antecedent diagnosis of autism,^5^ intellectual disability,^6^ Tourette syndrome/Tic disorder, ^7^ and attention-deficit/hyperactivity disorder (ADHD).^8^ If these associations are shown to be robust, of the neurodevelopmental conditions implicated in mortality risk, ADHD holds the largest potential for prevention given that it is the most common (global prevalence 3-7%^9^); seemingly increasing in occurrence;^10^ and its core symptoms of age-inappropriate levels of distraction, restlessness, and impulsivity are eminently treatable via prescription psychostimulants.^11^

A recent meta-analysis of eight non-overlapping published studies found a doubling of mortality risk in individuals affected by ADHD.^8^ The better-powered of the identified studies were generated from electronic linkage which has the particular advantages of capturing full-nation cohorts, having modest loss to follow-up, and providing precise risk signals.^12-15^ Such datasets also typically lack high resolution data on potential explanatory characteristics and this hampers understanding of the mechanisms via which ADHD may elevate the risk of premature mortality, and therefore when and how intervention might take place.

There may be at least four mechanistic pathways through which ADHD may be embodied in order to elevate mortality (figure 1). Educational challenges and the resulting socioeconomic disadvantage are well-established consequences of ADHD^16,17^ and both have been repeatedly linked to premature mortality.^18,19^ Co-morbid mental illness, particularly anxiety and depression, which, as described, are associated with excess mortality, ^1,2^ are markedly more prevalent in the ADHD-affected relative to those free of the disorder.^16,17^ Relatedly, self-medication via cigarette smoking and harmful levels of alcohol consumption in people with this neurodevelopmental condition^16,17^ – nicotine as a supplement/substitute for prescription psychostimulants, and ethanol as a suppressant for distress symptoms – raises the possibility of an elevated risk of death in ADHD that is mediated via these risk-taking behaviours.^20,21^ Lastly, ADHD has been connected to an array of subsequent adverse health events such as hypertension^22^ and diabetes^23^ for which the burden of premature death is raised.^24,25^

**Figure 1.**
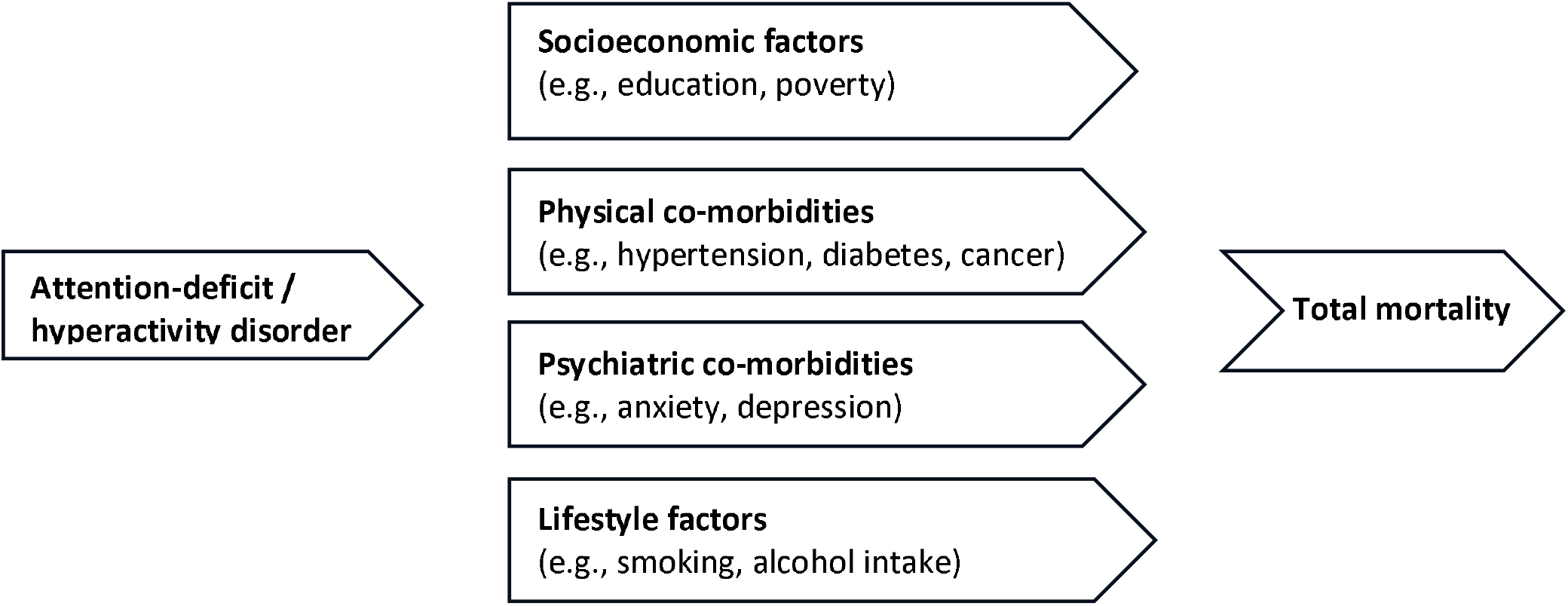
Hypothesized pathways connecting ADHD with all-cause mortality.

It is in this context that well-characterized, field-based cohort studies of the general population have particular utility. Using the US National Health Interview Survey (NHIS) we, first, add to the emerging evidence on mortality risk in people with ADHD; second, we examine the role of a wide array of risk factors in generating this relationship; third, we contextualize the role of ADHD by comparing any effect it may have on mortality against conventional risk factors; and, fourth, we explore the association of this neurodevelopmental disorder with specific causes of death, including cardiovascular disease and cancer, which may be driving the relationship with mortality but have thus far received little or no research attention.

## Methods

NHIS is a prospective cohort study based on annually-conducted, in-person, interviewer-administered household surveys of the health and associated behaviors of noninstitutionalized Americans who have been linked to death records.^26^ The baseline surveys, comprising around 30,000 adults aged >=18 years, have been conducted using a multistage, complex probabilistic design to ensure national representation. Data on ADHD diagnosis were collected in the 2007 (87% response) and 2012 (78%) surveys only.

Original data collection was approved by the National Center for Health Statistics Research Ethics Review Board. With the present publicly available data de-identified, further institutional review board approval was not required. Development of this manuscript followed the Strengthening the Reporting of Observational Studies in Epidemiology (STROBE) guidelines.^27^

### Assessment of ADHD

Study members were asked: “Have you ever been told by a doctor or other health professional that you had attention deficit hyperactivity disorder (ADHD) or attention deficit disorder (ADD)?”. This simple enquiry has been widely used in population-based studies^28^ and different lines of evidence support its validity. First, self-reported data from this question reveal a similar population prevalence to estimates based on clinical records in adults in the US,^29^ indicating convergent validity. Second, in prior analyses of the present dataset, people with a ADHD diagnosis were more likely to be male, ethnically white, and socioeconomically disadvantaged.^30^ That these differentials are well replicated, including in studies based on physician-verified diagnosis,^31^ suggests face validity. Third, in child and adolescent samples in NHIS, parents/carers were also administered the 25-item Strengths and Difficulties Questionnaire (SDQ) which captures ADHD symptoms. Study members who were ADHD positive based on these symptom scores (62%) were 9 times more likely to report a prior ADHD diagnosis relative to the SDQ ADHD negative group (7%).^32^ This suggests concurrent validity.

### Assessment of covariates

Of the demographic factors, self-declared race/ethnicity was categorized according to the US Office of Management and Budget:^33^ non-Hispanic White; non-Hispanic Black; Mexican American; other Hispanic; and ‘other’ race/ethnicity (comprises non-Hispanic Asian, American Indian/Alaska Native, Native Hawaiian/Other Pacific Islander, and people identifying as multi-racial). Of the socio-economic indicators, educational attainment was based on the highest level completed with response options collapsed into three groups: less than high school diploma (categories 0–3; lowest); some college (categories 4–7); college degree or higher (categories 8+). The ratio of family income to poverty was estimated by dividing household income by the US DHHS poverty threshold, with a ratio of ≥2 regarded as being advantageous.^34^

Lifestyle factors comprised health behaviors and body weight. A current smoker was defined as an individual who had reported smoking at least 100 cigarettes in their lifetime who was also presently smoking on some days or more. History of alcohol consumption was classified into lifetime abstinence, former, and current. People reporting no vigorous or moderate intensity physical activity per week were deemed sedentary. From self-reported height and weight, body mass index was calculated using the usual formula^35^ with 30 kg/m^2^ or more denoting obesity.

The six-item Kessler scale was used to assess depressive symptoms, with enquiries made about the occurrence of feelings of hopelessness, sadness, worthlessness, amongst others, in the preceding months.^36^ Responses were graded on a five point scale (‘none of the time [0]’ to ‘all of the time[4]’) and scores summed (range: 0 to 24). A total ≥13 indicated a higher probability of severe distress.^36^ For physical co-morbidities, enquiries were made about whether study members had been told by a doctor or health professional that they had hypertension, diabetes, stroke, heart disease, or cancer (yes/no).

### Ascertainment of mortality

Survey participants were linked to the National Death Index File; follow-up was from the date of interview to the date of death or 31 December 2019 – whichever came first. Our primary outcome was all-cause mortality. Underlying cause was coded according to the tenth revision of the International Classification of Disease (ICD)^37^ from which the secondary mortality outcomes of cardiovascular disease, all cancers combined, and chronic lower respiratory diseases were derived (for ICD codes, refer to supplemental table 4).

### Data analyses

As illustrated in figure 2, from an eligible sample of 57918 participants, we omitted those with missing data on ADHD diagnosis (N=81) and covariates (N=8521) plus failed linkage to mortality records (N=1390), resulting in an analytical sample of 52097. In our main analyses, we summarized the association of ADHD with mortality using Cox proportional hazards regression.^38^ After ascertaining that the association of ADHD with total mortality was near-identical in the 2007 (age, sex, race/ethnicity-adjusted hazard ratio [95% confidence interval]: 1.59 [1.08-2.34) and 2012 cohorts (1.59 [1.10-2.29]) (p-value for interaction = 0.975), data were pooled and adjusted for survey year. Hazard ratios and accompanying 95% confidence intervals were initially adjusted for demographic factors (age, sex, and race/ethnicity) and survey year (baseline hazard stratified). Adjusted cumulative mortality curves according to ADHD status were displayed using a flexible parametric Royston-Parmar model on the log cumulative hazard scale with 3 internal knots.^39^

**Figure 2.**
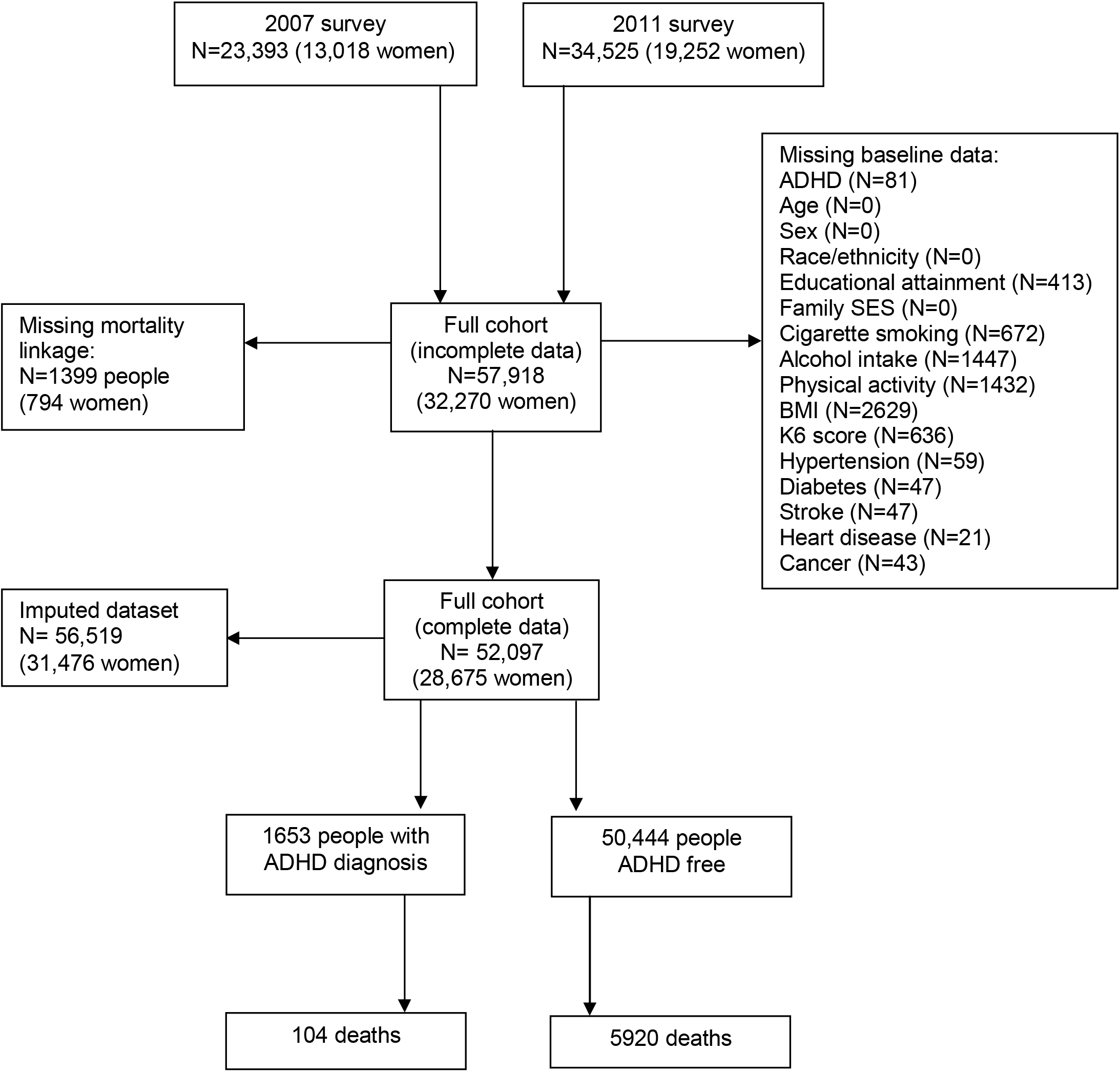
Flow of study members into the analytical samples (non-missing and imputed)

Based on a comparator model with adjustments for demographic factors and survey year, we then assessed the impact of separate control for a range of covariates according to theme: socioeconomic status (education, poverty), physical co-morbidities (cancer, hypertension etc), psychiatric co-morbidities (psychological distress), and lifestyle factors (body weight, cigarette smoking etc). We quantified any post-adjustment change in hazard ratio using this formula with the product expressed as percentage: (ln HR [Model 1] – ln HR [Model 2]) / ln HR [Model 1].^40,41^ Finally, multiple imputation by chained equations was used to explore the impact of missing covariates.^42^ All analyses were conducted using Stata (version 19.5) and R (version 4.5.2).

### Funding

This paper received no direct funding. GDB is supported by the UK Medical Research Council (MR/P023444/1) and the US National Institute on Aging (1R56AG052519-01, 1R01AG052519-01A1).

## Results

### Participant baseline characteristics according to ADHD diagnosis

In the analytical sample of 52097 people (28675 women), 1653 (3.2%) declared a baseline diagnosis of ADHD. In table 1 we show study member characteristics according to ADHD status (full distribution in supplemental 1). People with a ADHD diagnosis were more likely to be younger, male, and of white European ancestry than those who were free of the disorder. Of the socioeconomic factors, being in poorer circumstances was markedly more common in ADHD-affected study members, though educational disadvantage was the same as the ADHD-free group. While people reporting ADHD were twice as likely to smoke cigarettes and to drink alcohol, they were less liable to sedentary behavior. Owing to their lower baseline age, physical co-morbidities were markedly less common in people self-declaring ADHD, as evidenced by the prevalence of hypertension, diabetes, stroke, and cancer; psychological distress was markedly more common in the ADHD group, however.

**Table 1.** Study member baseline characteristics according to ADHD diagnosis.

|  | **ADHD** | **No ADHD** | **P-value** |
| --- | --- | --- | --- |
| Number of people | 1653 (3.2) | 50,444 (96.8) | - |
| **Survey year** |  |  |  |
| 2007 | 505 (39.0) | 19943 (48.0) | <0.001 |
| 2012 | 1148 (61.0) | 30501 (52.0) |  |
| **Demographic factors** |  |  |  |
| Age, mean (SD) | 33.9 (14.0) | 46.5 (17.7) | <0.001 |
| Male | 944 (63.6) | 22478 (48.5) | <0.001 |
| Non-Hispanic White | 1307 (84.3) | 30190 (67.9) | <0.001 |
| **Socioeconomic factors** |  |  |  |
| Less than high school education | 246 (14.5) | 8464 (14.6) | 0.090 |
| Socioeconomic disadvantage | 811 (41.6) | 19,126 (31.2) | <0.001 |
| **Lifestyle factors** |  |  |  |
| Current cigarette smoker | 618 (37.0) | 9332 (18.5) | <0.001 |
| Current alcohol drinker | 1206 (71.5) | 30935 (63.7) | <0.001 |
| Physically inactive | 445 (27.8) | 17965 (33.6) | <0.001 |
| Obese ($\geq 30$ kg/m<sup>2</sup>) | 466 (27.0) | 14218 (27.6) | 0.112 |
| **Psychological co-morbidity** |  |  |  |
| Distress, Mean (SD) | 5.2 (5.5) | 2.1 (3.6) | <0.001 |
| Severe Distress ($\geq 13$) | 228 (11.7) | 1457 (2.5) | <0.001 |
| **Physical comorbidities** |  |  |  |
| Hypertension | 415 (22.3) | 15964 (28.8) | <0.001 |
| Diabetes | 110 (5.3) | 4,04 (8.5) | <0.001 |
| Stroke | 39 (1.8) | 1540 (2.6) | 0.029 |
| Heart disease | 210 (12.1) | 6210 (11.4) | 0.525 |
| Cancer | 107 (5.3) | 4341 (8.1) | <0.001 |

Results are N (%) unless otherwise indicated.

### ADHD diagnosis and risk of all-cause mortality

A median 7.8 years of mortality surveillance (range: 7.3-12.3) gave rise to 6024 deaths from all-causes, with 104 occurring in people with a ADHD diagnosis (figure 2 and supplemental table 2). In survival analyses with basic adjustments (age, sex, ethnicity, and survey year), there was clear divergence in cumulative mortality according to baseline ADHD status, such that a markedly higher proportion of people with ADHD died than those without the disorder at all points during follow-up (figure 3). After the full surveillance period, this raised risk equated to a 58% higher mortality rate in ADHD-affected study members after the same adjustments (hazard ratio; 95% confidence interval: 1.58 [1.20-2.09]) (figure 4 and supplemental table 3).

**Figure 3.**
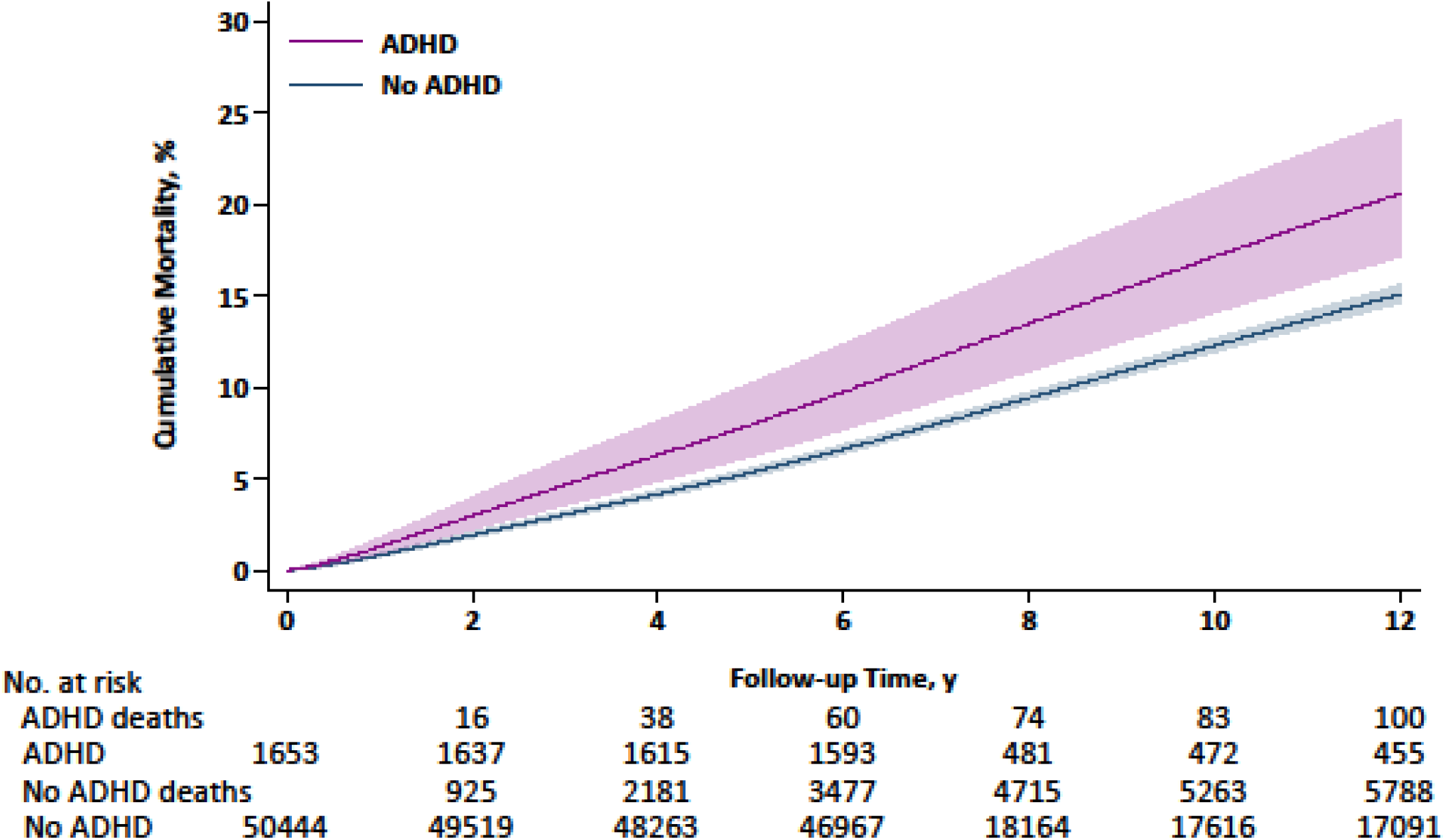
Survival of study members according to baseline ADHD diagnosis. Cumulative mortality curves are adjusted for demographic factors (age, sex, and race/ethnicity) and survey year. Shaded area indicates limits of 95% confidence intervals.

**Figure 4.**
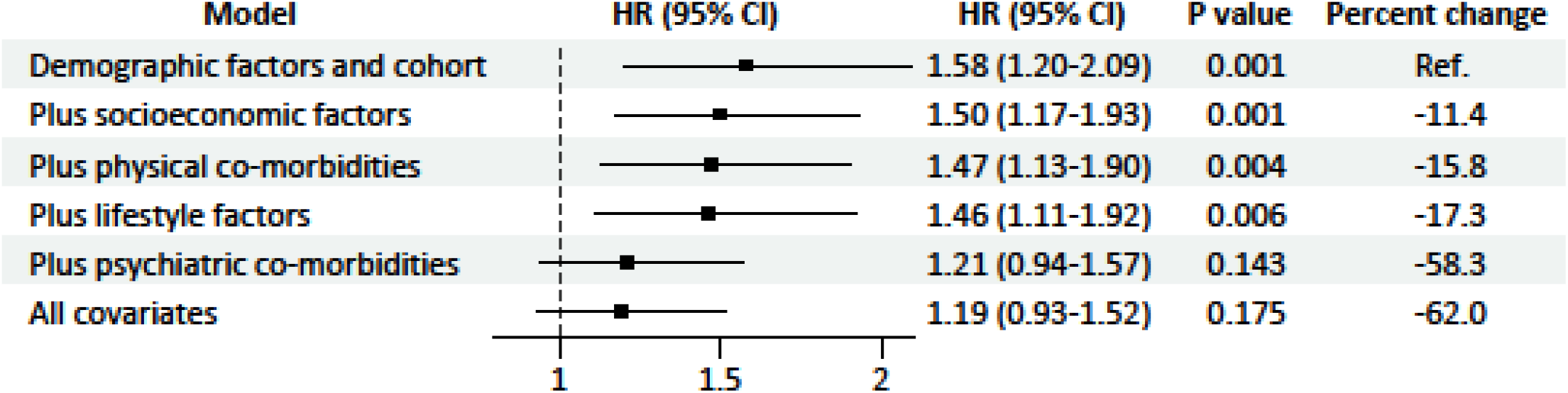
Association of baseline ADHD diagnosis with total mortality. Number of deaths according to ADHD diagnosis is given in supplemental table 2. Demographic factors comprise age, sex, and race/ethnicity; socioeconomic factors educational attainment and ratio of family income to poverty; physical co-morbidities hypertension, diabetes, stroke, heart disease, and cancer; lifestyle factors cigarette smoking, alcohol intake, physical activity, and body weight; and psychiatric co-morbidities psychological distress (anxiety/depression).

We then assessed the impact on the ADHD-death gradient, if any, of the explanatory factors depicted in figure 1. Relative to our comparator model, taking into account socioeconomic (11% attenuation), physical morbidities (15%), and lifestyle characteristics (17%) individually explained less than one-fifth of the ADHD– death relationship (figure 4). The most marked impact on the ADHD–death gradient was apparent after adjustment for psychological distress which explained 58% of the relationship and resulted in the raised mortality rate no longer being statistical significant at conventional levels (1.21 [0.94-1.57]). After multiple control for all collateral data, some of which were inter-correlated, the point estimate was little changed (1.19 [0.96-1.52]).

Next, to contextualize the magnitude of the association of ADHD with risk of total mortality, we compared it to well-established conventional predictors after basic adjustment (table 2). The hazard ratio for ADHD (1.58 [1.20-2.09]) was commensurate to that for basic educational attainment (1.77 [1.62-1.94]), poverty (1.66 [1.55-1.78]), hypertension (1.41 [1.32-1.51]), diabetes (1.71 [1.59-1.85]), heart disease (1.72 [1.61-1.83]), and cancer (1.33 [1.23-1.43]) but lower than cigarette smoking (2.51 [2.29-2.76]) and physical inactivity (2.05 [1.90-2.22]).

**Table 2.**
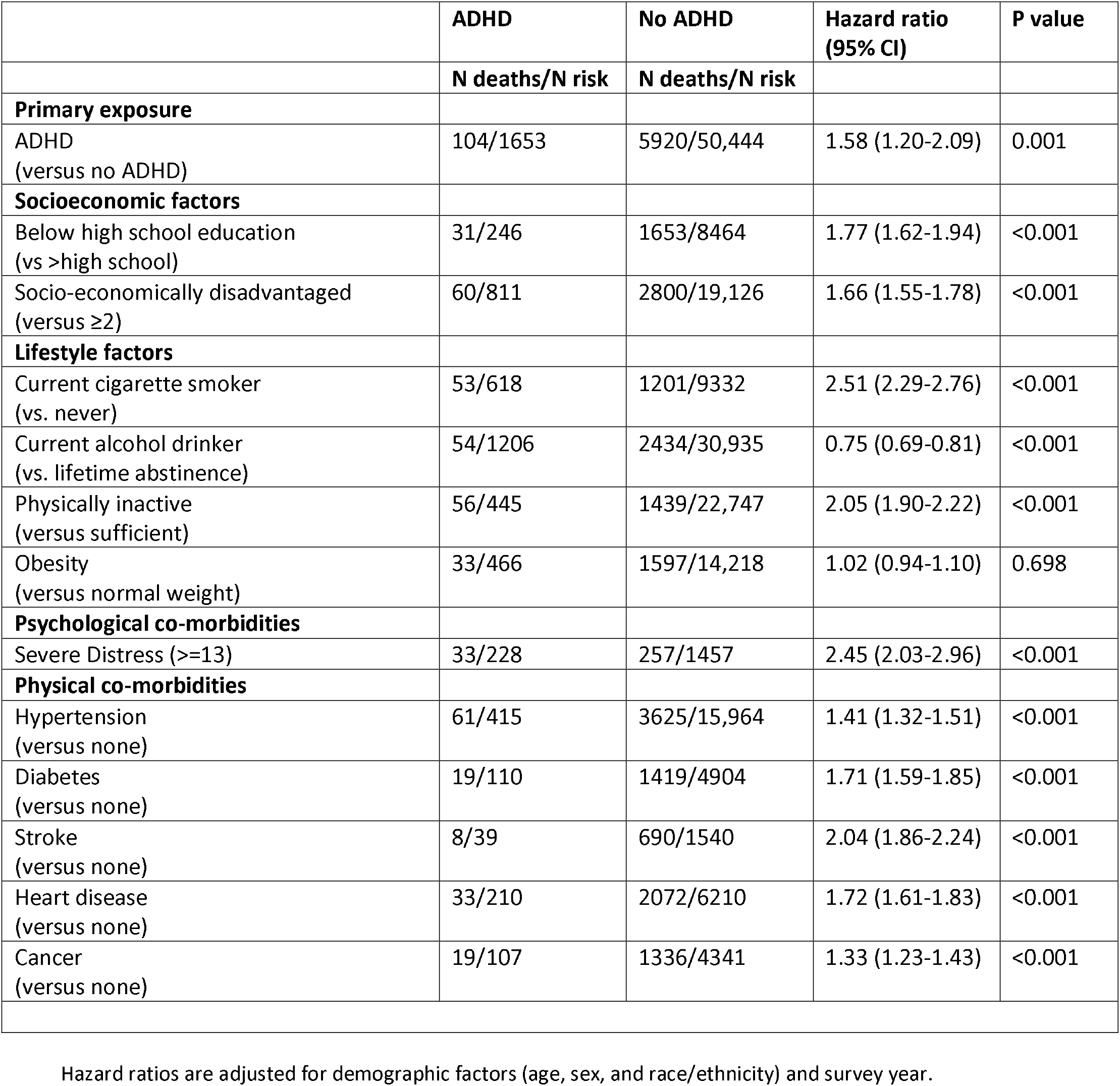
Association of baseline ADHD diagnosis and other modifiable risk factors with total mortality: Comparison of effect estimates.

|  | ADHD | No ADHD | Hazard ratio<br>(95% CI) | P value |
| --- | --- | --- | --- | --- |
|  | N deaths/N risk | N deaths/N risk |  |  |
| **Primary exposure** |  |  |  |  |
| ADHD<br>(versus no ADHD) | 104/1653 | 5920/50,444 | 1.58 (1.20-2.09) | 0.001 |
| **Socioeconomic factors** |  |  |  |  |
| Below high school education<br>(vs >high school) | 31/246 | 1653/8464 | 1.77 (1.62-1.94) | <0.001 |
| Socio-economically disadvantaged<br>(versus $\geq 2$) | 60/811 | 2800/19,126 | 1.66 (1.55-1.78) | <0.001 |
| **Lifestyle factors** |  |  |  |  |
| Current cigarette smoker<br>(vs. never) | 53/618 | 1201/9332 | 2.51 (2.29-2.76) | <0.001 |
| Current alcohol drinker<br>(vs. lifetime abstinence) | 54/1206 | 2434/30,935 | 0.75 (0.69-0.81) | <0.001 |
| Physically inactive<br>(versus sufficient) | 56/445 | 1439/22,747 | 2.05 (1.90-2.22) | <0.001 |
| Obesity<br>(versus normal weight) | 33/466 | 1597/14,218 | 1.02 (0.94-1.10) | 0.698 |
| **Psychological co-morbidities** |  |  |  |  |
| Severe Distress ($\geq 13$) | 33/228 | 257/1457 | 2.45 (2.03-2.96) | <0.001 |
| **Physical co-morbidities** |  |  |  |  |
| Hypertension<br>(versus none) | 61/415 | 3625/15,964 | 1.41 (1.32-1.51) | <0.001 |
| Diabetes<br>(versus none) | 19/110 | 1419/4904 | 1.71 (1.59-1.85) | <0.001 |
| Stroke<br>(versus none) | 8/39 | 690/1540 | 2.04 (1.86-2.24) | <0.001 |
| Heart disease<br>(versus none) | 33/210 | 2072/6210 | 1.72 (1.61-1.83) | <0.001 |
| Cancer<br>(versus none) | 19/107 | 1336/4341 | 1.33 (1.23-1.43) | <0.001 |

Hazard ratios are adjusted for demographic factors (age, sex, and race/ethnicity) and survey year.

### ADHD diagnosis and risk of cause-specific mortality

To examine which causes of death might be generating the association between ADHD and total mortality, we carried out survival analyses for specific adverse health outcomes. With the exception of residual deaths, the number of cause-specific fatalities in the ADHD group was low (supplemental table 2) and this diminished statistical precision as evidenced by the wide confidence intervals in some analyses (supplemental table 4). This notwithstanding, rates of mortality in people with ADHD were elevated relative to those without the condition for all specific causes of death, although not at conventional levels of statistical significance and the relationship for cardiovascular disease was particularly weak. The only exception was for residual deaths for which there was around a doubling of risk in ADHD-affected people (1.88 [1.33-2.64]). After adjustment for covariates, the pattern of attenuation was similar to that for total mortality whereby the largest explanatory impact was again apparent for psychological distress.

### Sensitivity analyses

We carried out some sensitivity analyses. First, as described, there was inevitably some missing data at study induction which was highest for the lifestyle factors (figure 2) and in our main analyses we used a non-missing dataset (N=52,097). Omitting study members with partial data may introduce some selection bias. Following multiple imputation, we generated a new analytical sample (N=56,519) and survival analyses produced essentially the same hazard ratios as those reported in the main analyses (supplemental table 5). Second, the Kessler distress scale contains an enquiry about frequency of feeling restless/fidgety which is one of the diagnostic characteristics for hyperactivity in ADHD.^43^ Statistical adjustment for distress (anxiety and depression) measured using this inventory could therefore have led to an underestimation of the true strength of the ADHD–mortality relation. We therefore substituted the distress data for self-report of a health professional diagnosis of anxiety and/or depression (enquiries were the same as for hypertension etc). Adjusting separately for these diagnoses (1.26 [0.97-1.63]; p-value 0.080) in comparison to the Kessler scale as described in the main analyses (1.21 [0.94, 1.57[) did not alter our conclusion about the ADHD–death relationship.

## Discussion

Our analyses revealed a series of new findings. First, while the markedly raised rate of all-cause mortality in people with self-declared ADHD was partially ascribed to socioeconomic disadvantage, physical co-morbidities, and lifestyle factors, symptoms of psychological distress exerted most explanatory power. This suggests that the influence of ADHD on mortality was largely embodied via anxiety and depression in the present study. Second, ADHD appeared to have similar predictive capacity for total mortality relative to several more established risk indices, several of which are known to be causal factors in the occurrence of premature mortality (e.g., hypertension, diabetes). Finally, disaggregating total mortality into specific causes of death revealed little association with common chronic disease outcomes such as all cancers and cardiovascular disease, though robust effects were seen for ADHD and the heterogeneous residual category of causes of death.

### Comparison with existing studies

Although comparatively few such studies exist relative to other psychiatric exposures such as serious mental illness, there is a suggestion of elevated mortality rates in the ADHD-affected.^8^ Hitherto, explanations for this association have been scant given the rarity of high resolution covariate data in the registry-based cohort studies on which these findings are largely based, however, there is a suggestion that socioeconomic disadvantage and mental illness, which have been captured in the most comprehensive data linkages, have a partial mediating role.^15,44^ Importantly, we are unaware of studies which have explored the impact of lifestyle factors such as cigarette smoking, physical inactivity, body weight, and alcohol intake as we have here.

Studies aggregating chronic diseases into a single death-from-natural-causes endpoint typically report no association with antecedent ADHD.^15^ For specific disease outcomes, investigators on a cross-sectional study found an association between ADHD and self-reported CVD^30^ that was subsequently verified in the only prospective cohort conducted to date.^45^ In this, a higher incidence of CVD episodes requiring hospitalization relative to disorder-free controls was reported.^45^ While we found no evidence of a correlation between ADHD and deaths ascribed to cardiovascular disease, these analyses, alongside those for cancer and respiratory disease, which revealed moderately raised but non-significant hazard ratios after basic adjustment, were hampered by a low number of events in people with this neurodevelopmental condition. Our models for residual causes of death were better powered and the pattern of association closely resembled that for total mortality. A portion of these events would have comprised external causes of death such as intentional (e.g., suicide) and unintentional (e.g., road traffic accidents) injury – both health endpoints that have been connected to earlier ADHD diagnosis in epidemiological studies.^46,47^

### Study strengths and limitations

The present study is well-characterized for explanatory factors, has modest loss to follow-up, and allows a prospective examination of the association between ADHD and adverse health outcomes. Our work is of course not without its limitations, however, and these provide direction for future research. First, we did not have data on specific ADHD symptoms in order to explore their separate effects on mortality. It is plausible for instance that the greater volume of energy expenditure seen in people who are hyperactive^48^ may be protective against premature mortality, while, as described, inattention to health messaging, such as the avoidance of weight gain, could elevate risk. Second, relatedly, having data on symptom load would have facilitated a dose-response analyses and its application would be particularly useful in people who are sub-diagnostic but nonetheless have a greater burden of ADHD symptoms. With this group being considerably more populous than those meeting a ADHD diagnostic threshold, even a moderately elevated mortality risk will have a higher population attributable risk. Third, even after multivariable adjustment for all relevant covariates, there remained some excess mortality in ADHD-affected people. Selected risk factors such as hypertension and diabetes are somewhat hidden conditions such that direct measurement in biomedical surveys of general population samples reveal new cases.^49^ These data were not captured herein and nor was information on loneliness and social isolation which may also hold some explanatory power.

In conclusion, in the present study, the influence of ADHD on mortality risk appears to be largely embodied via lifestyle factors, physical comorbidities, and, particularly, mental illness. While supporting evidence is required, our findings point to the possibility that modification of these characteristics via pharmacological treatment and behavioral intervention may have utility in the avoidance of premature mortality in people with ADHD.

## Data Availability

Data acquisition: Deidentified NHIS data are publicly available (https://www.cdc.gov/nchs/nhis/documentation/index.html). Restricted data can be sought under application.

## Supplemental file

Hazard ratios are adjusted for demographic factors (age, sex, and race/ethnicity) and survey year.

**Supplemental table 1.**
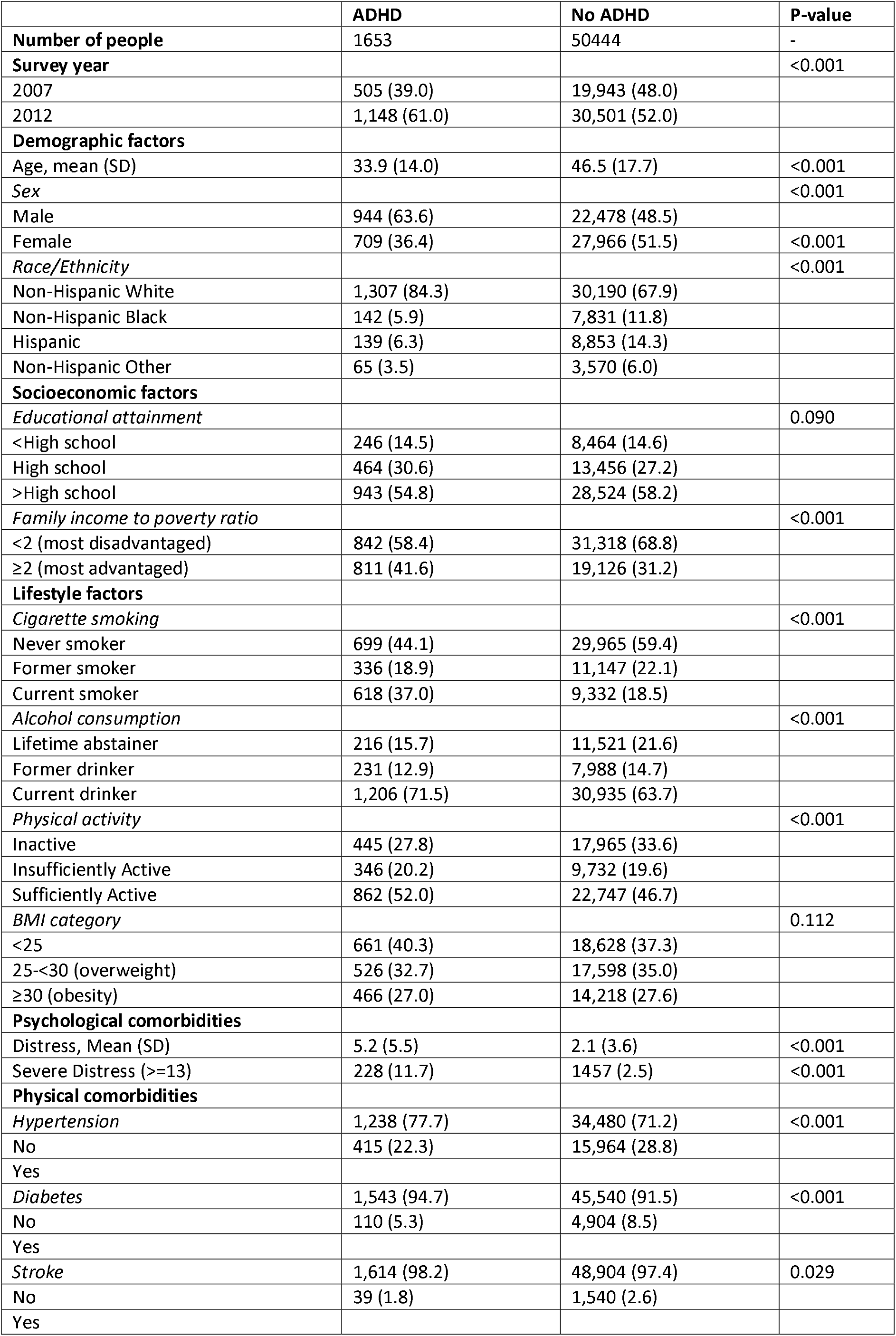
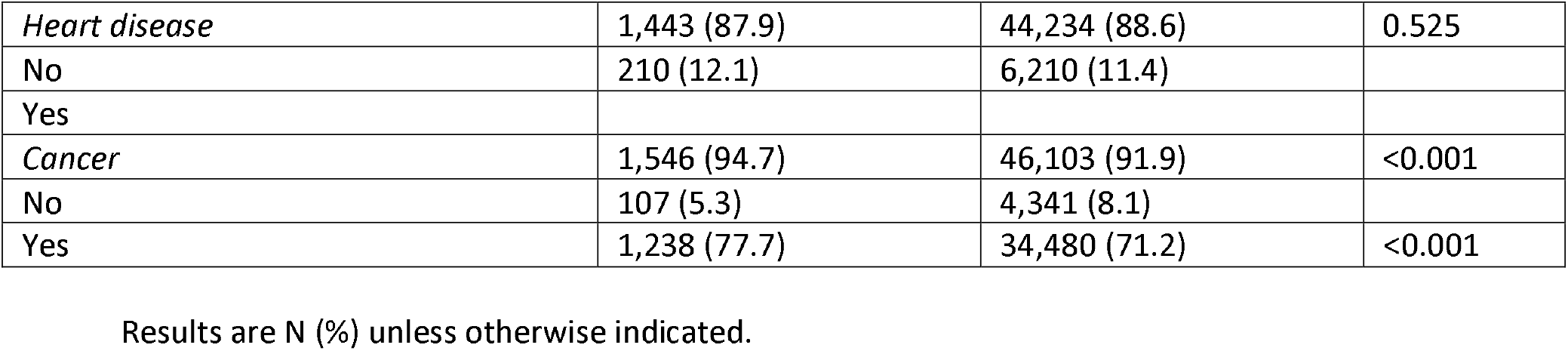
Study member baseline characteristics according to ADHD diagnosis.

**Supplemental table 2.** Study member follow-up characteristics according to baseline ADHD diagnosis.

|  | **ADHD** | **No ADHD** | **P-value** |
| --- | --- | --- | --- |
| Number of people | 1653 | 50444 |  |
| Median follow-up (IQR), y | 7.75 (7.25-12.25) | 7.75 (7.25-12.25) | - |
| Person-years at risk | 74,353,888 | 2,056,822,183 | - |
| All-cause mortality | 104 (5.3) | 5,920 (10.3) | <0.001 |
| Cardiovascular disease mortality | 19 (0.9) | 1,772 (3.0) | <0.001 |
| Cancer mortality | 28 (1.5) | 1,367 (2.6) | <0.001 |
| Chronic lower respiratory diseases | 6 (0.3) | 373 (0.7) | 0.018 |
| All other causes (residual) | 51 (2.6) | 2,408 (4.1) | <0.001 |

Results are N (%) unless otherwise indicated.

**Supplemental Table 3.** Association of baseline ADHD diagnosis with total mortality.

| Model | Hazard ratio<br>(95% confidence<br>interval) | P value | Percent<br>change |
| --- | --- | --- | --- |
| Demographic factors and cohort | 1.58 (1.20-2.09) | 0.001 | Ref. |
| Plus socioeconomic factors | 1.50 (1.17-1.93) | 0.001 | -11.4 |
| Plus physical co-morbidities | 1.47 (1.13-1.90) | 0.004 | -15.8 |
| Plus lifestyle factors | 1.46 (1.11-1.92) | 0.006 | -17.3 |
| Plus psychiatric co-morbidities | 1.21 (0.94-1.57) | 0.143 | -58.3 |
| All covariates | 1.19 (0.93-1.52) | 0.175 | -62.0 |

Hazard ratios are ordered according to degree of attention. Demographic factors comprise age, sex, and race/ethnicity; socioeconomic factors educational attainment and ratio of family income to poverty; physical co-morbidities hypertension, diabetes, stroke, heart disease, and cancer; lifestyle factors cigarette smoking, alcohol intake, physical activity, and body weight; and psychiatric co-morbidities psychological distress (anxiety/depression). Percent change quantifies the impact of serial adjustment on the HR from the basic model.

**Supplemental Table 4.** Association of baseline ADHD diagnosis with cause-specific mortality.

|  | Hazard ratio<br>(95% confidence<br>interval) | P value | Percent<br>change |
| --- | --- | --- | --- |
| **Cardiovascular disease** |  |  |  |
| Demographic factors and cohort | 1.13 (0.62-2.04) | 0.691 | Ref. |
| Plus socioeconomic factors | 1.10 (0.63-1.93) | 0.739 | -22 |
| Plus physical co-morbidities | 1.06 (0.62-1.84) | 0.821 | -52.3 |
| Plus lifestyle factors | 1.06 (0.58-1.92) | 0.854 | -52.3 |
| Plus psychiatric co-morbidities | 0.91 (0.52-1.61) | 0.751 | -177.2 |
| All covariates | 0.94 (0.54-1.63) | 0.816 | -150.6 |
| **All cancers** |  |  |  |
| Demographic factors and cohort | 1.51 (0.98-2.34) | 0.064 | Ref. |
| Plus socioeconomic factors | 1.38 (0.55-3.49) | 0.493 | -21.8 |
| Plus physical co-morbidities | 1.37 (0.54-3.47) | 0.507 | -23.6 |
| Plus lifestyle factors | 1.32 (0.51-3.43) | 0.569 | -32.6 |
| Plus psychiatric co-morbidities | 0.95 (0.38-2.35) | 0.907 | -112.4 |
| All covariates | 1.24 (0.78-1.97) | 0.353 | -47.8 |
| **Chronic lower respiratory diseases** |  |  |  |
| Demographic factors and cohort | 1.49 (0.58-3.82) | 0.400 | Ref. |
| Plus socioeconomic factors | 1.38 (0.55-3.49) | 0.493 | -21.8 |
| Plus physical co-morbidities | 1.37 (0.54-3.47) | 0.507 | -21.1 |
| Plus lifestyle factors | 1.32 (0.51-3.43) | 0.569 | -30.4 |
| Plus psychiatric co-morbidities | 0.95 (0.38-2.35) | 0.907 | -112.9 |
| All covariates | 1.00 (0.41-2.45) | >0.999 | -100.0 |
| **Other causes (residual)** |  |  |  |
| Demographic factors and cohort | 1.88 (1.33-2.64) | <0.001 | Ref. |
| Plus socioeconomic factors | 1.73 (1.25-2.40) | 0.001 | -13.2 |
| Plus physical co-morbidities | 1.78 (1.28-2.47) | 0.001 | -8.7 |
| Plus lifestyle factors | 1.75 (1.25-2.45) | 0.001 | -11.4 |
| Plus psychiatric co-morbidities | 1.34 (0.95-1.87) | 0.092 | -53.6 |
| All covariates | 1.30 (0.93-1.82) | 0.121 | -58.4 |

Demographic factors comprise age, sex, and race/ethnicity; socioeconomic factors educational attainment and ratio of family income to poverty; physical co-morbidities hypertension, diabetes, stroke, heart disease, and cancer; lifestyle factors cigarette smoking, alcohol intake, physical activity, and body weight; and psychiatric co-morbidities psychological distress (anxiety/depression). Number of deaths for each outcome according to ADHD diagnosis is given in supplemental table 2. Percent change quantifies the impact of serial adjustment on the HR from the basic model.
Cardiovascular disease comprises ischemic heart disease (ICD 10 codes: I00-I09, I11, I13, I20-I51) and cerebrovascular diseases (I60-I69). All cancers combined were denoted by C00-C97, and for chronic lower respiratory diseases, we used codes J40-J47.

**Supplemental Table 5.** Association of baseline ADHD diagnosis with total mortality: Multiple imputation for missing data.

| Model | Hazard ratio<br>(95% confidence<br>interval) | P value | Percent<br>change |
| --- | --- | --- | --- |
| Demographic factors and cohort | 1.59 (1.22-2.07) | 0.001 | Ref. |
| Plus socioeconomic factors | 1.51 (1.19-1.92) | 0.001 | -11.1 |
| Plus physical co-morbidities | 1.46 (1.15-1.87) | 0.002 | -18.4 |
| Plus lifestyle factors | 1.46 (1.12-1.89) | 0.005 | -18.4 |
| Plus psychiatric co-morbidities | 1.22 (0.95-1.56) | 0.121 | -57.1 |
| All covariates | 1.18 (0.93-1.50) | 0.162 | -64.3 |

Demographic factors comprise age, sex, and race/ethnicity; socioeconomic factors educational attainment and ratio of family income to poverty; physical co-morbidities hypertension, diabetes, stroke, heart disease, and cancer; lifestyle factors cigarette smoking, alcohol intake, physical activity, and body weight; and psychiatric co-morbidities psychological distress (anxiety/depression). Number of deaths according to ADHD diagnosis is given in supplemental table 2. Percent change quantifies the impact of serial adjustment on the HR from the basic model.

